# Neuroimmune transcriptome changes in brains of psychiatric and neurological disorder patients

**DOI:** 10.1101/2022.02.14.22269692

**Authors:** Yu Chen, Jiacheng Dai, Longfei Tang, Tatiana Mikhailova, Qiuman Liang, Miao Li, Jiaqi Zhou, Cynthia Weickert, Chao Chen, Chunyu Liu

## Abstract

Neuroinflammation has been implicated in multiple brain disorders but the extent and the magnitude of change in immune-related genes (IRGs) across distinct brain disorders has not been directly compared. We curated 1,275 IRGs and investigated their expression changes in 2,467 postmortem brains of controls and patients with six major brain disorders, including schizophrenia (SCZ), bipolar disorder (BD), autism spectrum disorder (ASD), major depressive disorder (MDD), Alzheimer’s disease (AD), and Parkinson’s disease (PD). More than 60% of the IRGs had significantly altered expression in at least one of the six disorders. The differentially expressed immune-related genes (dIRGs) shared across disorders were mainly related to innate immunity. Moreover, we systematically evaluated sex, tissue, and cell type for immune alterations in different neuropsychiatric disorders. Co-expression networks revealed that neuroimmune systems interacted with neuronal-systems, both of which contribute to the risk of disorders. However, only a few genes with expression changes have also been identified as containing risk variants of genome-wide association studies. The transcriptome alterations at gene and network levels may clarify the immune-related pathophysiology and redefine neuropsychiatric and neurological disorders.

**One-Sentence Summary:** The brain transcriptome of six neurological and psychiatric disorders showed signature changes in genes related to immunity.

## Introduction

Multiple lines of evidence support the notion that the immune system is involved in major “brain disorders,” including psychiatric disorders such as schizophrenia (SCZ) (*1*), bipolar disorder (BD) (*2*), and major depressive disorder (MDD)(*3*), brain development disorders such as autism spectrum disorder (ASD)(*4*), and neurodegenerative diseases such as Alzheimer’s disease (AD)(*5*), and Parkinson’s disease (PD)(*6*). Patients with these brain diseases share deficits in cognition, blunted mood, restricted sociability and abnormal behavior to various degrees. Transcriptome studies have identified expression alterations of immune-related genes (IRGs) in postmortem brains of AD(*7*), PD(*8*), ASD(*9*), SCZ(*10-14*) and BD(*10*) separately. Cross-disorder transcriptomic studies further highlighted changes in IRGs(*15, 16*). At the protein level, several peripheral cytokines showed reproducible disease-specific changes in a meta-analysis (*17*). Since brain dysfunction is considered the major cause of these disorders, studying immune gene expression changes in patient brains may reveal mechanistic connections between immune system genes and brain dysfunction. Most previous studies were limited to the analysis of individual disorders. There is no comprehensive comparison of the pattern and extent of inflammation-related changes in terms of immune constructs (subnetworks), neuro-immune interaction, genetic contribution, and relationship between diseases.

Neuroinflammation, an immune response taking place within the central nervous system (CNS), can be activated by psychological stress, aging, infection, trauma, ischemia, and toxins (*18, 19*). It is regulated by sex (*20*), tissue type (*21*) and genetics (*22*), many of which are known disease risk factors for both psychiatric and neurological diseases. The primary function of neuroinflammation is to maintain brain homeostasis through protection (*23*) and repair (*24*). Abnormal neuroinflammation activation could lead to dysregulation of mood (*25*), social behaviors (*26*), and cognitive abilities (*27*). Offspring who were fetuses when their mothers’ immune system was activated (MIA) showed dopaminergic hyperfunction (*28*), cognitive impairment (*29*), and behavioral abnormalities (*30*) as adults. Alternatively, acute and chronic neuroinflammation in adulthood can also alter cognition and behavior(*31, 32*). In animal models, both adult and developmental maternal immune activation in the periphery can lead to increases in pro-inflammatory cytokines in the brain, similar to what is found in humans with major mental illness(*14, 33*).

Previous studies identified immune gene dysregulations in brains of patients with several major brain disorders. For example, Gandal et al.(*16*) found that up-regulated genes and isoforms in SCZ, BD, and ASD were enriched in pathways such as inflammatory response and response to cytokines. One brain co-expression module up-regulated specifically in MDD was enriched for genes of cytokine-cytokine interactions, and hormone activity pathways*(15)*. The association of neurological diseases such as AD and PD with IRGs has also been reported(*7, 34*). These studies examined the changes of immune system as a whole without going into details of specific subnetworks, the disease signature, or genetic versus environmental contribution.

We hypothesize that expression changes of specific subsets of IRGs constitute part of the transcriptome signatures that distinguishes diseases. Since tissue specificity, sex and genetics all could influence such transcriptome signatures, we analyzed their effects. Furthermore, we expect that neurological diseases and psychiatric disorders bear transcriptomic changes that may help to address how similar immunological mechanisms lead to distinct brain disorders. The current boundary between neurological diseases and psychiatric disorders is primarily the presence of known pathology. Neurological diseases have more robust histological changes while psychiatric disorders have more subtle subcellular changes. Nonetheless, pathology evidence is always a subject to be revised with new research.

To investigate immune-related signatures of transcriptome dysregulation in brains of six neurological and psychiatric disorders, we studied a selected list of 1,275 genes known to be associated with neuroinflammation and interrogated their expression across disorders. We collected and analyzed existing transcriptome data of 2,467 postmortem brain samples from donors with AD(*35-37*), ASD(*38-41*), BD(*38, 42-45*), MDD(*44, 46*), PD(*47-50*), SCZ(*38, 42-45, 51, 52*) and healthy controls (CTL). We identified the differentially expressed IRGs shared across disorders or specific to each disorder, and their related coexpression modules (Fig. S1). These genes and their networks and pathways provided important insight into how immunity may contribute to the risk of these neurological and psychiatric disorders, with a potential to refine disease classification.

## Results

### Expressions of immune-related genes were altered in brain disorders

We collected 1,789 IRGs from curated databases including Comparative Toxicogenomics Database(*53*), ImmPort(*54*), ImmunomeDB(*55*), InnateDB(*56*), ImmuneSigDB(*57*), Gene Ontology database with immune annotation(*58*), KEGG database with immune annotation(*59*), as well as additional literature reviews (*60-62*), followed by filtering based on expression profile in the human brains (see Table. S**1**). We compiled 23 transcriptomic datasets of multiple brain disorders from the Gene Expression Omnibus (GEO), ArrayExpress, or from the authors directly (see Table. S**1**). In total, we collected transcriptome data of 2,467 postmortem brain samples from subjects with AD (n = 340 individuals), ASD (n = 103), BD (n = 188), MDD (n =87), PD (n = 97), SCZ (n = 474) and CTL (n = 1,178). We used the data sets derived from microarray (n=1007) as our discovery set and data sets derived from RNA-Seq as an independent replication set (Table. S1). Out of the 1,789 selected IRGs, 1,275 (71%) were detected across all the microarray data. The IRG curation procedure is summarized in a Preferred Reporting Items for Systematic Reviews and Meta-Analyses (PRISMA) workflow (Fig. S2). **Table S2** lists the 1,275 detected IRGs categorized by their reference databases and pathway annotation programs.

After preprocessing the microarray data (methods and materials), we conducted a whole transcriptome differential expression analysis for each disorder using a linear mixed-effects model that accounted for sample overlap across studies. Filtering the differentially expressed genes (DEGs, FDR<0.05) for the 1,275 IRGs, we identified dIRGs in each of the six brain disorders (number of dIRGs in descending order of transcript identified as differentially expressed; AD: 638, ASD: 275, SCZ: 220, PD: 97, BD: 58, MDD: 27, Fig. 1A). We used a conservative genome-wide threshold instead of study-wide threshold for significance so that we could further evaluate the relative enrichment of IRGs in all the DEGs under the same significance criteria. The dIRGs were significantly over-represented in the DEGs across disorders (OR>2, adjust.p < 0.05, Table S3). The enrichment of IRGs in the DEGs supports the reported immune gene dysfunction in transcriptome of these six disorders, highlighting its importance.

**Fig. 1.**
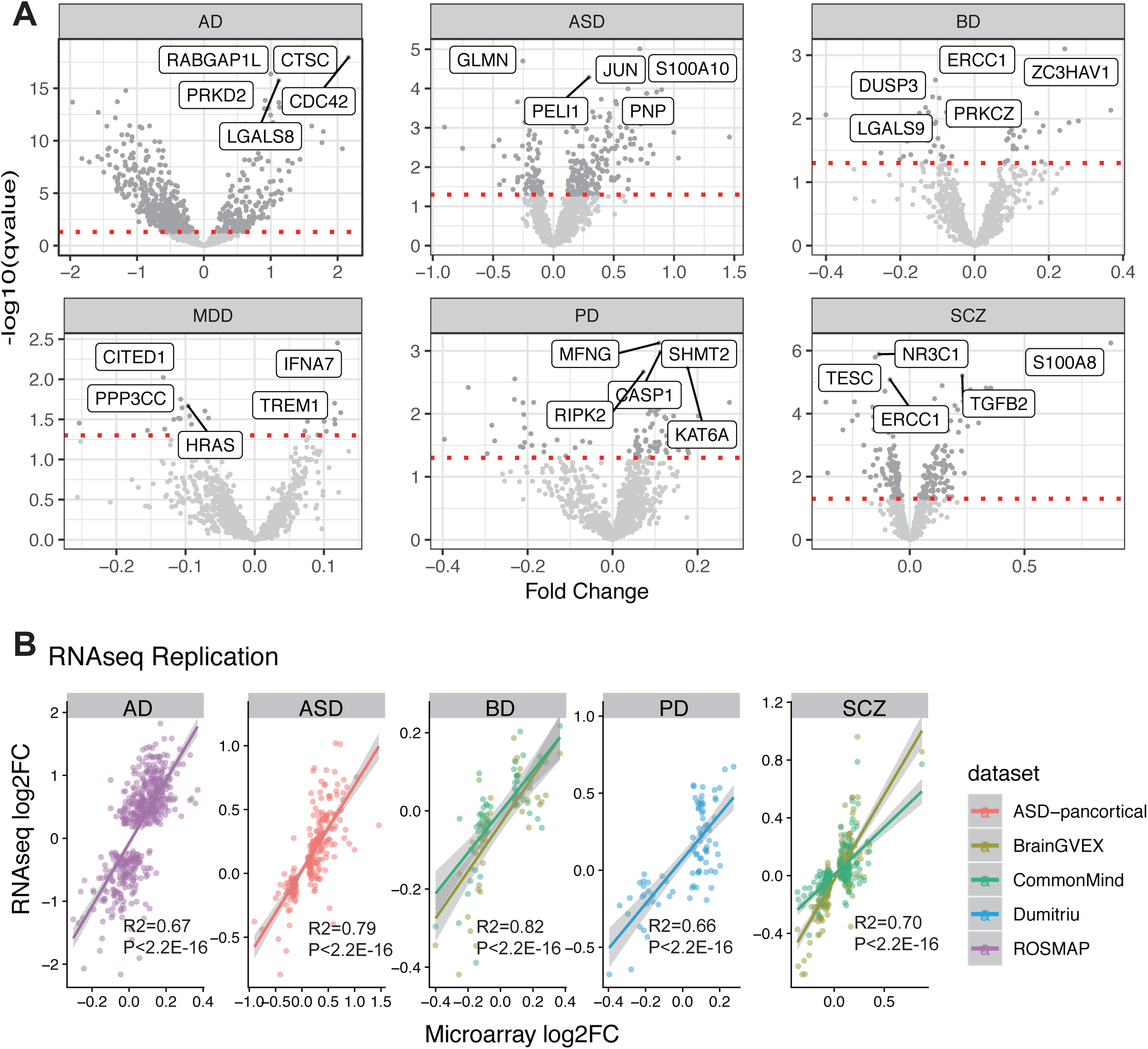
Differential expression of immune genes in six disorders. A. Volcano plot for each disorder. B. Effect size correlation between microarray data and RNA-seq data.

To replicate the findings, we used the independent RNA-seq datasets and processed data as detailed in the Methods and Materials. We observed a significant overlap of dIRGs between discovery and replicate datasets (Table S4). AD data achieved the highest replication rate (56%) while BD achieved the lowest (19%). More importantly, we observed high concordance of effect sizes of case-control fold change between the microarray and RNA-seq results for all IRGs (R2 > 0.66, p.value < 2.2E-16, Fig. 1B, Table S4).

### The changes of IRGs clustered by disorders and sex- and tissue-specific effects

We used hierarchical clustering based on the correlations of the fold changes of all the detected IRGs among different brain disorders, resulting in two distinct groups, one containing all psychiatric disorders (BD, SCZ, and MDD) and another containing mainly neurological disorders (ASD, AD, and PD) (Fig. 2A, B). The fold changes of IRGs were highly correlated between SCZ and BD (Spearman’s r = 0.75, p.value <0.001). When comparing the groups of psychiatric disorders and neurological disorders, we found a higher effect size (larger fold-change) in the expression of inflammatory related genes in neurological disorders than in psychiatric disorders (t-test p.value < 2.2E-16, Fig. S4). RNA-seq data replicated the observation (Fig. S4) that larger immune-related dysregulation was present in neurological disorders than in psychiatric disorders.

**Fig. 2.**
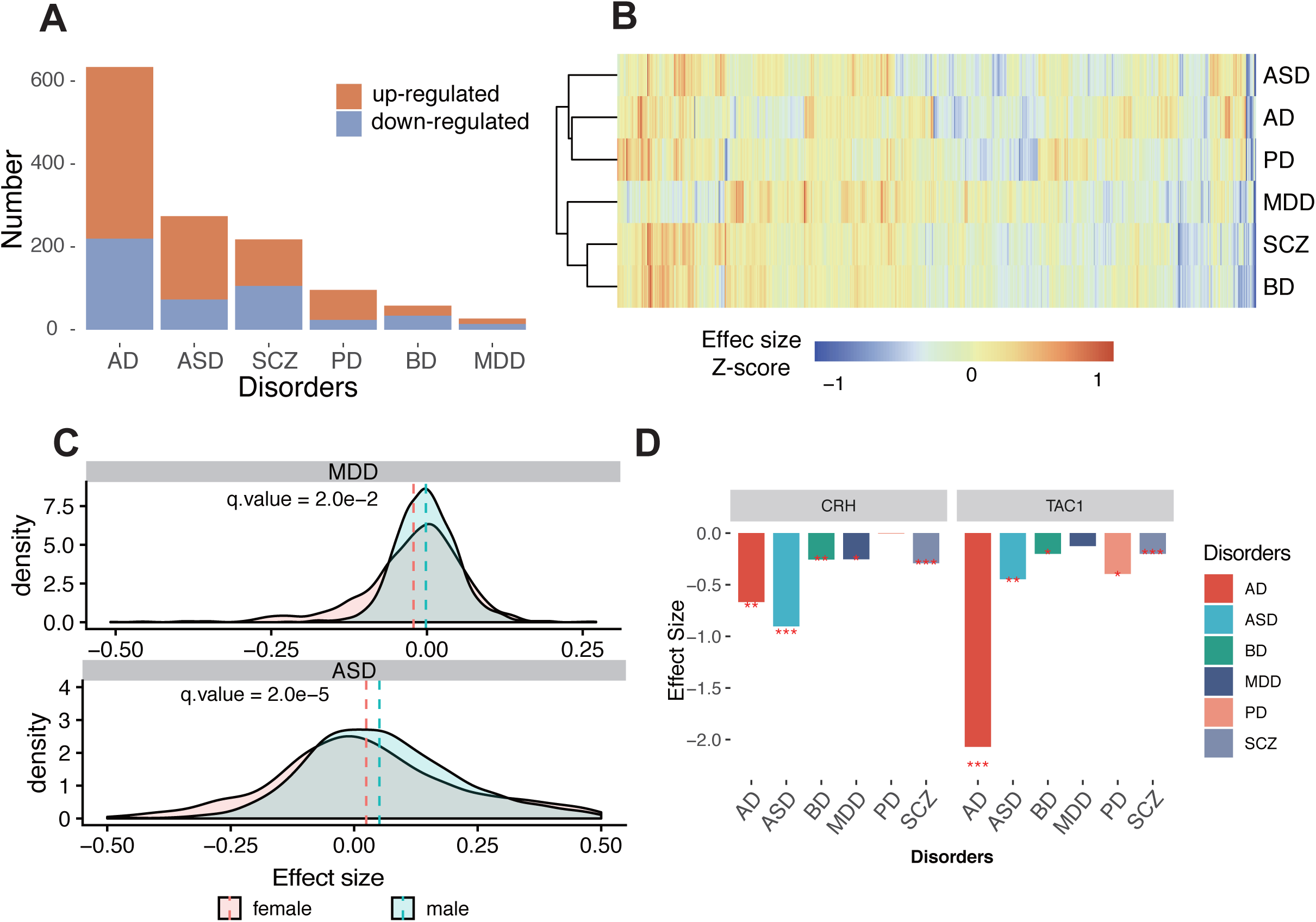
Comparison of the effect size of differentially expressed IRGs among neuropsychiatric disease pairs. A. Numbers of up-regulated and downregulated dIRGs in the six disorders. Red represents up-regulated dIRGs, blue represents down-regulated dIRGs. B. Cluster tree of scaled effect size for all disorders based on 1275 IRGs for their fold changes. C. Significant sex differences by effect size in ASD and MDD. D. dIRGs shared across disorders: CRH and TAC1. *: fdr q.value<0.05; **fdr q.value<0.01; ***fdr q.value<0.001

To test effects of sex on immune-related dysregulation, we recomputed dIRGs with the samples partitioned by sex. Comparing the effect sizes of the IRGs between male and female subgroups, we found significant sex differences in ASD and MDD (Fig. 2C, Table. S5; p_ASD_ = 0.003; p_MDD_ = 4E-6), but not in other diseases. The IRGs showed larger magnitude of change in male ASD than in female ASD relative to corresponding controls, while the situation was the opposite for MDD with females having larger changes than males.

To investigate tissue specificity of IRG dysregulation, or more specifically, whether alterations of IRGs in the brain can be reflected in blood, we calculated the changes of IRGs in blood datasets of these six disorders (Table S1). The correlation of IRGs’ effect size showed negligible concordance (R^2^ from -0.24 to 0.11, P-value>0.05, Fig. S5), indicating that the majority of the changes of IRGs in the blood and brain do not overlap, implying distinct origins and/or cellular mechanisms. However, we still identified a few dIRGs showing consistent changes in brain and blood (Table S6), such as S100A8 in SCZ. These genes may serve as candidates of disease peripheral biomarkers, which warrants a thorough investigation.

### Innate immune genes are the most shared changes across all brain disorders

Comparing the overlap of dIRGs across disorders, 26% of IRGs were dIRGs in two or more disorders (shared dIRGs, Fig. 3 A, B). While we found alterations of both adaptive and innate IRGs in each disease (Fig. S6, Table S3), 68% of the shared dIRGs were classified as genes involving innate immune functions. To avoid bias caused by the number of IRGs in the two categories, we further calculated the enrichment of dIRG and found that they were significantly enriched in only innate IRGs (Fisher Exact Test OR > 2, qvalue <0.05). In the RNA-seq replicate data, there was a better replication rates in innate IRGs than in adaptive IRGs (Fig. S6, Table S4).

**Fig. 3.**
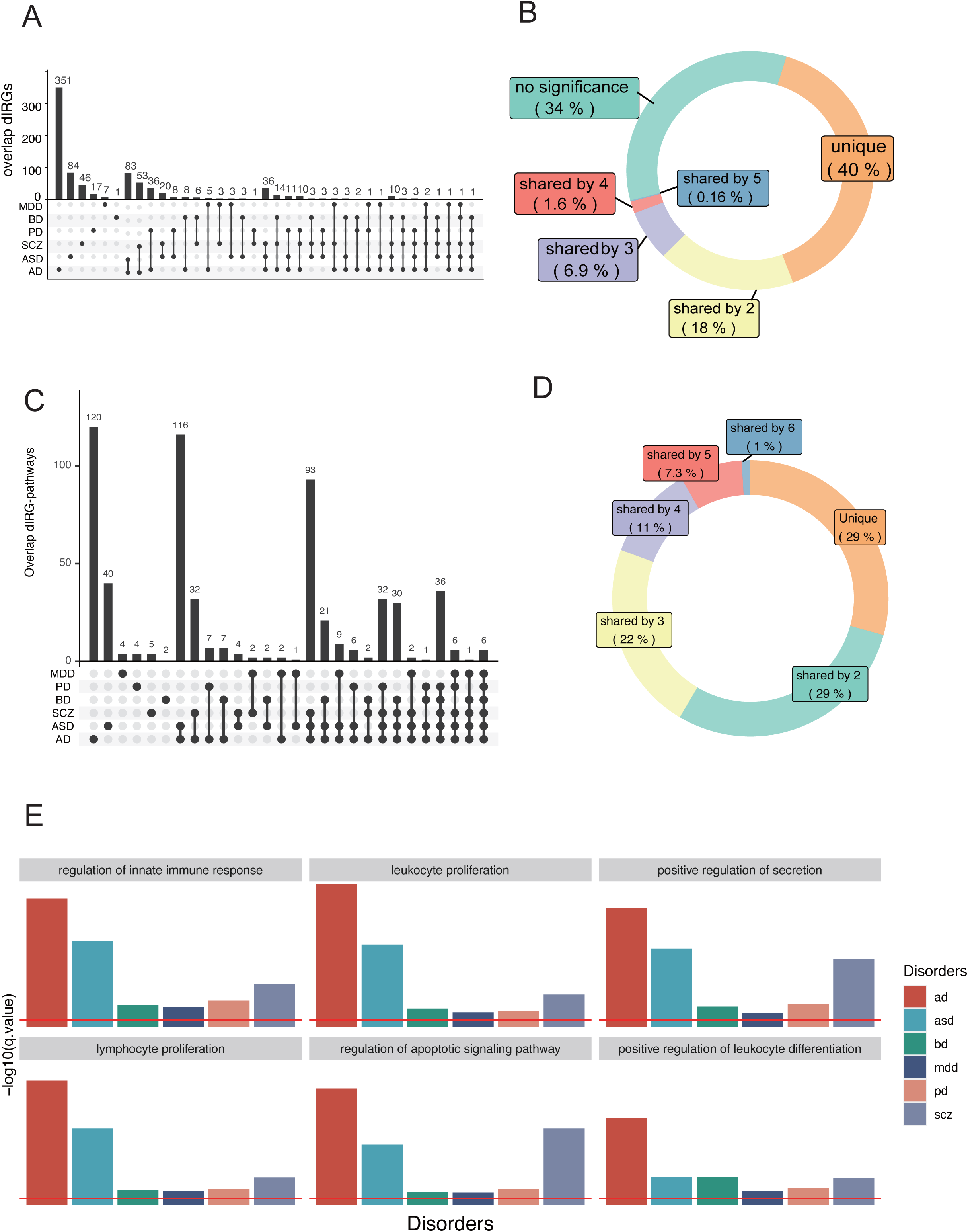
Comparing dIRG-associated function across disorders. A. UpSet plot of dIRGs overlap between pairs of disorders. Dark cells and lines indicate that the set participates in the intersection. B. The doughnut chart shows the percentages of different IRGs types. C. UpSet plot of differential immune pathways overlap. The black dots and the black line show the overlapping dIRG-pathways between pairs of disorders. Cells that are dark indicate that it participates in the intersection. D. The doughnut chart shows the percentages of overlapping dIRG-pathways. E. Gene ontology enrichment analysis results of six pathways shared by dIRGs of all six disorders.

The two most shared dIRGs are Corticotropin-releasing hormone (CRH) and Tachykinin Precursor 1 (TAC1), which were differentially expressed in five of the six diseases (Fig. 2D). They both involve innate immunity according to the databases we used and literature (*53-56, 58, 59*). CRH was downregulated in five of the six disorders; the exception was PD. CRH can regulate innate immune activation with neurotensin (NT), stimulating mast cells, endothelia, and microglia (57). TAC1 was down-regulated in five of the six disorders, the exception being MDD. TAC1 encodes four products of substance P, which can alter the immune functions of activated microglia and astrocytes (*63*). Independent RNA-seq data confirmed both CRH and TAC1 findings. These transcripts are also neuromodulators and have action on neurons so they have roles in addition to immune functions.

To identify specific immune pathways that dIRGs are involved in, we further tested the enrichment of the dIRGs in specific immune functions. We found six dIRGs-enriched pathways shared by all six disorders. “Regulation of innate immune response (GO:0045088)” was one of the six pathways (q.value< 0.05, Table. S7, Fig. 3E). When a subset of the innate immune genes as defined by the GO database was used as input instead of all IRGs, hierarchical clustering resulted in the same clusters of psychiatric vs. neurological diseases (Fig. S7). This indicated that even though immune dysfunction is widespread in the six disorders, signature patterns of the subset innate immune genes are sufficient to differentiate neurological from psychiatric disorders.

### Immune-related coexpression modules (IRMs) shared by diseases were related to brain development and aging

To determine if neuroimmunity works in silos or cooperates with other functions, we used robust weighted gene coexpression network analysis (rWGCNA)(*64*) to construct immune-related co-expression networks on the whole transcriptome. We identified 16 brain co-expression networks shared across disorders (Fig. 4A, Fig. S7) after adjusting the batch covariate. Three of the 16 networks were enriched for IRGs and were called immune-related co-expression modules (Fig. 4B, C; IRM4, IRG enrichment q.value = 2.15E-2; IRM12, q.value = 1.13E-04; IRM14, q.value = 9.29E-12). These three IRMs were significantly associated with at least two disorders in the same direction (Fig. 4D). The Eigengene (hub gene) of IRM4 was negatively associated with all disorders (Fig. 4D). IRM4 was enriched for neuron markers (Fig. 4E). Two of the modules (IRM12 and IRM14) were enriched with microglia and endothelial marker genes (Fig. 4E), respectively, and were both positively associated with IRGs in AD and ASD (Fig. 4D). AD had the strongest association with all three IRMs (Fig. 4D). The endothelial-related IRM14 and neuron-related IRM4 were both enriched for tissue development (q.value= 6.69E-5, Fig. 4F, Table S8) and neuron development (q.value= 6.49E-11, Fig. 4F, Table S8). Additionally, the IRM4 was significantly enriched for late fetal cortical markers (q.value= 1.921E-07, Table. S8).

**Fig. 4.**
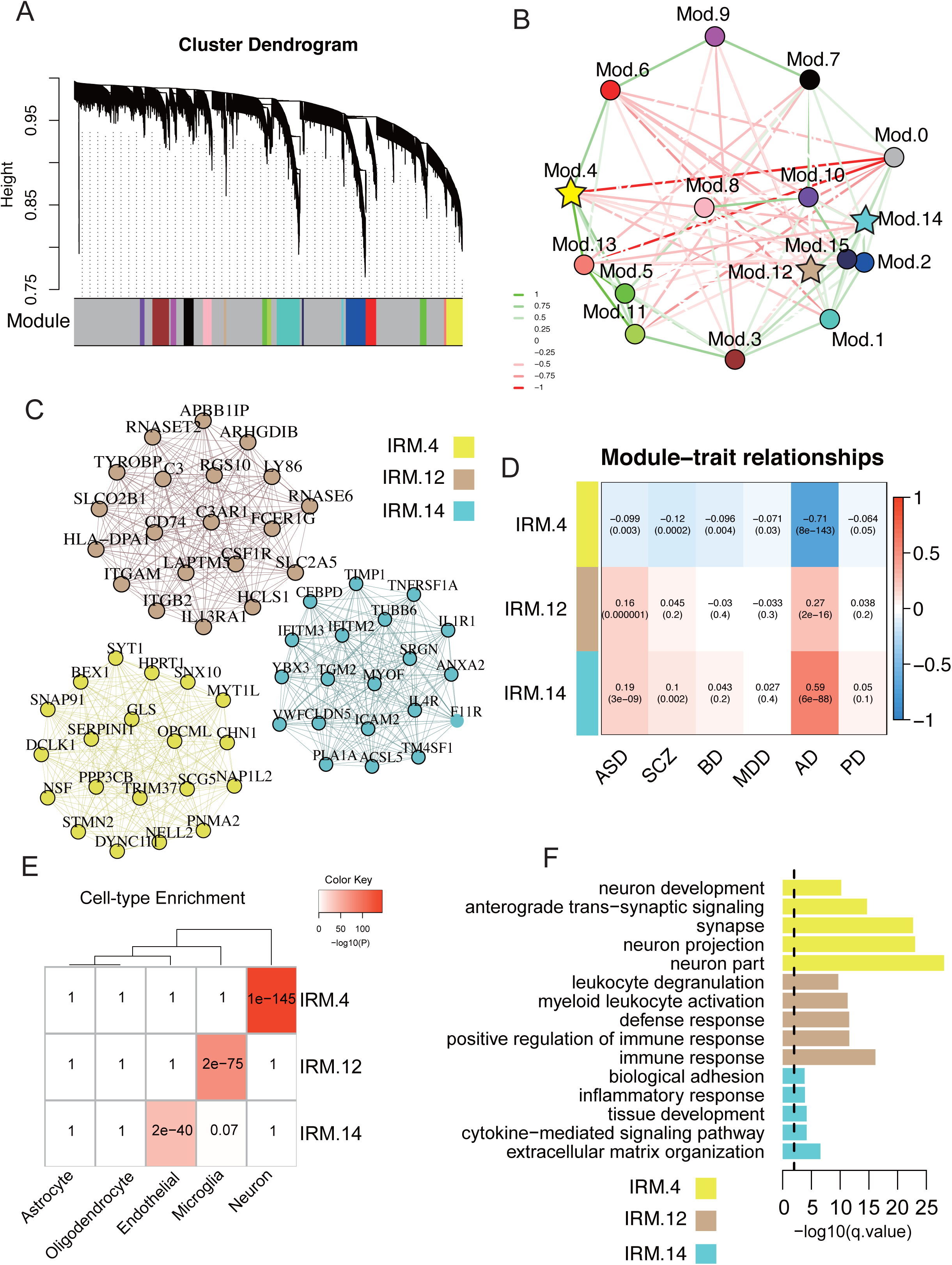
Shared immune-related coexpression modules. **A**. Robust gene dendrogram obtained by WGCNA. **B**. The multidimensional scaling plot demonstrates the relationship between modules. Modules highlighted by stars are enriched in immune genes (enrichment q-value < 0.05). Edges are weighted by the strength of correlation between eigengenes of modules. **C**. The top 20 hub genes are plotted for the three IRM4, IRM12, and IRM14. A complete list of genes’ module membership (kME) is provided in data Table S8. Edges are weighted by the strength of the correlation between genes. **D**. Relationships of module eigengenes and diseases. Numbers in the table report the correlations of the corresponding module eigengenes and traits, with the p.values printed below the correlation coefficients r values. **E**. Cell marker enrichment of shared IRMs. **F**. Enrichment of the shared IRMs in pathways. Yellow: IRM4, Tan: IRM12, Cyan: IRM14

We also assessed the influence of age on the IRMs and found that the age trajectories of these IRMs in cases had distinct patterns across disorders (Fig. S7D), which further illustrated the disease-specific temporal dynamics of these IRMs. For example, the neuron-related IRM-4 genes are continuously up-regulated in AD, with inverted U-shaped curve peaking at age ∼80, while PD shows a continuous downward trend in the same age range.

### Disease-specific IRMs in AD, ASD, and PD imply distinct biological processes

We also searched for disease-specific IRMs for each disorder. We used rWGCNA to construct brain co-expression networks in the brains of each disorder and of controls, then compared them against each other to identify **disease-specific IRMs** (Fig.5A). Based on preservation results of one disease versus controls and against all other diseases (Fig. 5B, z-summary < 10), as well as immune gene enrichment results (Table S9; enrichment q.value < 0.05), we identified six disease-specific IRMs, including one for AD, three for ASD, and two for PD. We did not detect disease-specific IRMs for SCZ, BD, or MDD, which are considered psychiatric disorders.

**Fig. 5.**
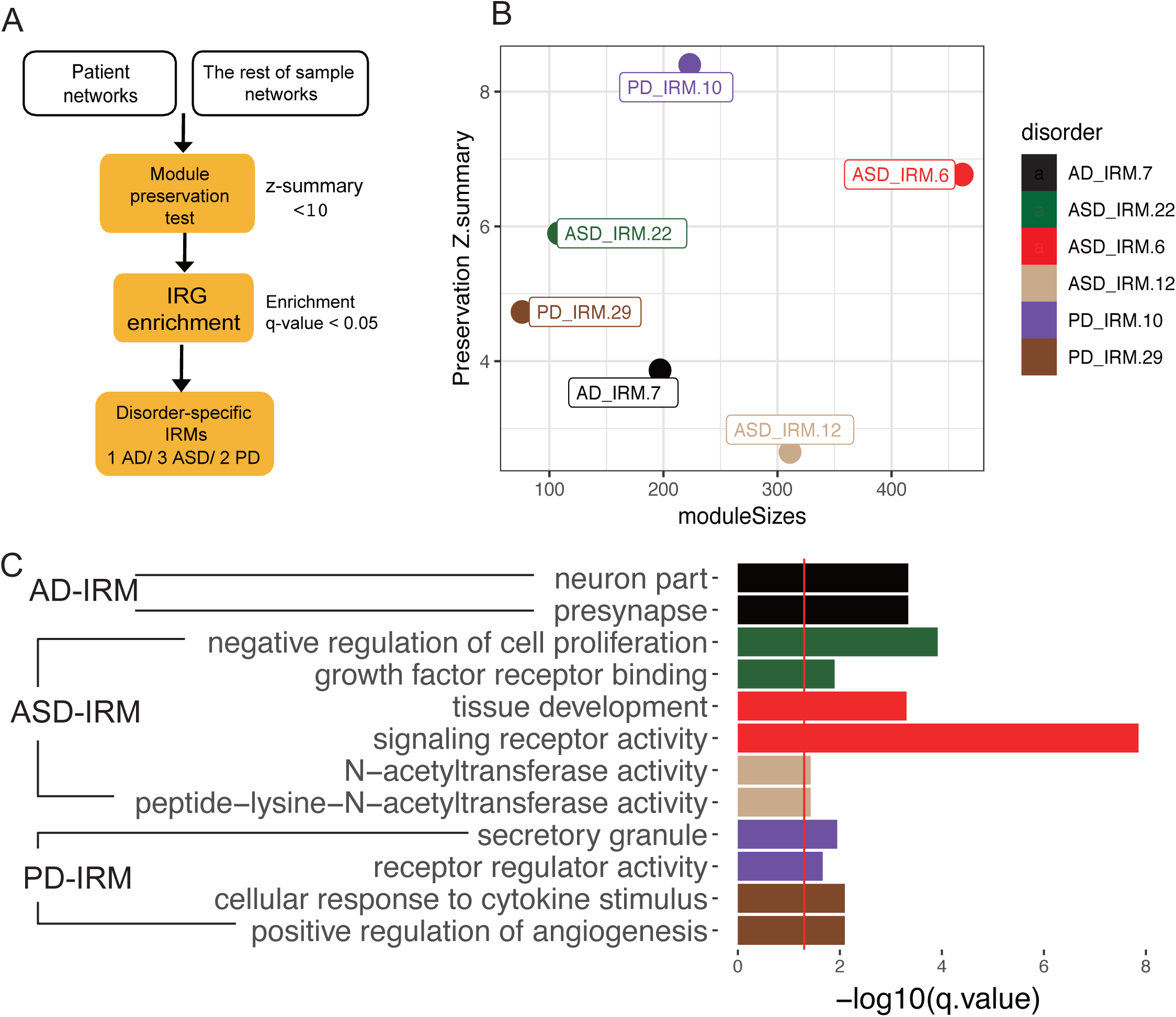
Disease-specific coexpression modules. **A**. Workflow for identifying disease-specific IRMs. **B**. Module preservation plot of disease-specific IRMs. The median rank and Zsummary statistics of module preservation of disorder modules in background modules (y-axis) vs. module size (x-axis). **C**. Pathway enrichment of disease-specific immune modules.

The disease-specific IRMs were enriched for various functions (Fig. 5C, Table S9). The AD-specific IRM was enriched for neuron part (GO:0097458, q.value= 4.57E-4) and presynapse (GO:0098793, q.value = 4.57E-4). The PD-specific IRM was enriched for positive regulation of angiogenesis (GO:0045766, q.value = 9.65E-06) and secretory granule (GO:0030141, q.value= 6.31E-06). The ASD-specific IRMs were enriched for developmental biological processes such as negative regulation of cell proliferation (GO:0008285, q.value= 1.21E-4) and growth factor receptor binding (GO:0070851, q.value = 1.27E-02).

### Common SNPs have a modest contribution to neuroimmune changes

We used two different approaches to further explore whether the altered IRGs in brain disorders were influenced by common SNPs.We examined sharing between dIRGs and disease GWAS signal genes and the correlation relationships between pairs of disorders for their changes of dIRGs and disease heritability calculated by GWAS.

We tested whether dIRGs, dIRG-enriched pathways, and IRMs were enriched for disease GWAS signals. We found a few dIRGs overlapping with genes that are located in GWAS loci for all six diseases, with AD having the largest number of genes (35 genes), and SCZ the second-largest (15 genes), including IL6R in AD, C4A in the SCZ. (Table S10). A few dIRG-related pathways were significantly enriched in AD and SCZ GWAS signals and survived multiple testing corrections (Table S10). Among them, the amyloid precursor protein catabolic process was enriched in AD GWAS signals (q.value= 3.9E-7). The leukocyte apoptotic process was enriched in SCZ GWAS signals (q.value= 0.03). Only IRM-4 was enriched in SCZ GWAS signals (Table S10).

When two disorders have similar genetic risks, will they have similar changes of IRGs? In other words, is the genetic similarity between two disorders reflected by the similarity of expression changes of IRGs? We assessed the relationship between the effect-size correlation of dIRGs and the genetic correlation from the same pairs of disorders(15). The genetic correlation was obtained from the Brainstorm Consortium(*65*). Putting all the pairs of disorders together, we obtained a correlation of these two kinds of correlations, modest correlation by r value, but insignificant by p value (Fig. S8, Pearson’ r = 0.46, p.value = 0.08). Though there was a small sample size of pairs of disorders (N=15) in this analysis, the analysis suggested that genetic factors had a minor contribution to general brain disorders through affecting the IRGs. This analysis captures collective contributions from all six disorders and cannot resolve individual contributions.

## Discussion

Our study focused on the neuroimmune changes represented by gene expression in multiple neuropsychiatric and neurological disorders. We used transcriptome data of more than 2,000 brains from healthy controls and patients of six major brain disorders. By studying individual IRGs and related pathways and coexpression networks, we found that brain disorders have both shared and disease-specific immune-related changes. In addition, we evaluated the effects of biological factors such as tissue, sex, age, and cell type. We came up with four major findings of the neuroimmune system in brains of different neuropsychiatric disorders: 1) the innate immune system carries more alterations than the adaptive immune systems in the six disorders; 2) the altered immune systems interact with other biological pathways and networks contributing to the risk of disorders; 3) common SNPs have a limited contribution to immune-related disease risks, suggesting theenvironmental contribution may be substantial; and 4) the expression profiles of dIRGs, particularly that of innate immune genes, group neurodevelopment disorder ASD with neurological diseases (AD and PD) instead of with psychiatric disorders (BD, MDD, and SCZ) Dysregulation of the innate immune system is a common denominator for all six brain disorders. We found that more than half of the shared dIRGs and dIRG-enriched pathways were related to the innate immune system. The two most shared dIRGs, TAC1 and CRH, have known effects on innate immune activation(*66, 67*). Both genes were downregulated in patient brains. Additionally, TLR1/2 mediates microglial activity, which could contribute to neuronal death through the release of inflammatory mediators(*68*). Furthermore, innate immunity is critical in maintaining homeostasis in the brain. For example, the innate immune system has been reported to function in the CNS’s resilience(*69*) and in synaptic pruning throughout brain growth(*70*). When homeostasis is disrupted, the abnormal innate immunity may impact a wide range of brain functions(*71*).

Concerning tissue specificity, our results indicated that there are huge differences in immune transcriptomes between blood and brain in patients with brain disorders, highlighting the importance of studying immune systems in brain. This inconsistency can be caused by the blood-brain barrier, which separates the CNS from the peripheral circulation. CNS and blood have independent responses to immune-related insults(*72*).

Microglia are highlighted in the immune changes in brains of AD and ASD in this study. Microglia is the major cell type participating in the brain’s immune system. Our analyses showed that the IRM12 coexpression module was enriched for microglia genes and associated with inflammatory transcriptional change in AD and ASD but not the other four diseases. Does this suggest that microglial dysfunction contributes more to AD and ASD than to the other disorders? The PsychENCODE study showed the microglial module upregulated in ASD and downregulated in SCZ and BD(*16*), but the fold changes in SCZ and BD were much smaller than that in ASD (Fig 7.B in original paper(*16*)). Larger sample size may be needed to detect microglia contribution to other disorders such as SCZ and BD.

Sex contributes to the disease-related immune changes too. Our results revealed sex-bias dysregulation of IRGs in brains of ASD and MDD but not in other disorders. These two disorders are known to have sex differences in prevalence(*73-75*). Previous studies also have suggested that sex differences in stress-related neuroinflammation might account for the overall sex bias in stress-linked psychiatric disorders, including female bias in MDD(*76*) and male bias in ASD(*77*). We did not observe sex-biased IRGs in other diseases with known sex-biased prevalence, such as SCZ and AD suggesting that sex differences in SCZ and AD may not involve IRG changes.

The dIRGs are expected to be enriched for immune-related pathways since we selected genes from immune systems. Therefore, it is more interesting to learn how the neuroimmune system cooperates with other brain-related biological processes to influence the disease risk. Our coexpression network analyses suggested that IRGs’ changes in patients’ brains were involved in developmental processes. Three brain universal IRMs were also enriched with functions related to development in neurons, microglia, and endothelia. Among them, IRM4 is of particular interest for connecting neuron, immune system, and development to all six disorders tested in this study. The contribution of neuro-immune-development to SCZ and ASD is well accepted. Previous studies discovered the role of the immune system in the development of the CNS(*69, 78, 79*). Abnormal immune activation during brain development can cause behavioral and neurochemical abnormalities relevant to disorders(*80-84*). One additional disease-specific module, ASD-IRM6, is also associated with development, further implicating the importance of development in ASD risk. The connections between AD, PD, and neuro-immune-development through IRM4 remain unclear, as they are late-onset neurodegenerative diseases. The unique age trajectories of these IRM4 in AD patients suggested aging in the immune system was involved for the same set of genes.

Our results showed how immune system dysregulation may influence gene expression of the networked other non-immune genes and contribute to the pathology of these diseases specifically. Six disease-specific IRMs were detected in AD, ASD, and PD, showing that several functions of the immune-related networks also involved in corresponding disorders such as presynaptic-related AD-IRM and Growth factor receptors-related ASD-IRMs. Presynaptic proteins are essential for synaptic function and are related to cognitive impairments in AD(*85*). Growth factor receptors(*86*) and N-acetylcysteine(*87*) are involved in the etiology of ASD. Secretogranin may be a pivotal component of the neuroendocrine pathway and play an essential role in neuronal communication and neurotransmitter release in PD (*88*). Furthermore, the immune system has been found to regulate presynaptic proteins(*89*), EGFR(*90*), and secretogranin(*88*). Our results indicate that alterations of the immune network can be disease-specific, affecting specific coexpression networks and driving distinct risk of each disorder.

Debate exists over whether neuroinflammatory alterations in disorders are affected by common SNPs(*91, 92*) or by the environment(*93, 94*). This study offered support for both arguments. On the genetic risk side, we did not detect statistically significant correlation between overall genetic risk and IRG changes between pairs of disorders. This IRG-subset result is in contrast with the results on whole transcriptome(15) where a significant correlation with Spearman’s ρ =0.79 was reported. Such difference suggests the environmental contributions to the immune-related risks in these disorders. dIRGs were not significantly enriched in any disease GWAS signals. A number of dIRGs are also GWAS signals such as IL6R in AD and C4A in SCZ. IL6R has been identified as a strong candidate gene of AD with both genetic and transcriptome supports(*95*). C4A is a well-known risk gene which was identified by SCZ GWAS and is up-regulated in SCZ (effect size=0.2, qvalue<0.05). Several dIRG-related pathways and coexpression modules were enriched in GWAS signals of neurological diseases AD and SCZ, respectively. Enrichment for AD was particularly strong, involving the amyloid precursor protein catabolic process in AD. A previous study identified this pathway, which is under genetic control in AD(*96*). The genetic connection with immunity detected here is certainly indirect. We did not detect any significant enrichment of dIRG in GWASs of ASD, PD, BD, and MDD. The genetic contribution to neuroinflammation may be more relevant to AD and SCZ than the other four disorders. Our interpretation is that the immune contributions to the risks of all these disorders were mostly related to the environment.

The environmental contribution to neuropsychiatric disease risk is strongly implicated through immunity. Our data suggests that these brain diseases are related to stress, an environmental factor. CRH is one of the most shared dIRGs across disorders. CRH has the core function of controlling the release of stress hormones. Studies have reported the relationship between immunity and stress(*97, 98*) and showed patients with brain disorders had decreased cortisol responses to social stressors(*77*). BD-specific immune-related pathway is reflected in genes changed in a mouse stress model, further emphasizing the contribution of stress to BD.

To our surprise, neurodevelopment disorder ASD was grouped with neurological diseases (AD and PD) instead of with psychiatric disorders (BD, MDD, and SCZ) according to the changes of IRGs, particularly innate immune genes. Hierarchical clustering analysis based on the effect size of IRGs placed the presumed psychiatric disorder ASD with other neurological diseases. Previous studies have reported that ASD patients exhibited more neurological and immunological problems(*99-102*) compared to healthy people and to other brain disorders. As more etiologies are uncovered, the traditional classification of these diseases is increasingly challenged(*93*). Furthermore, we found that dIRGs change more in neurological diseases (AD, PD, and ASD) than in the psychiatric disorders (BD, SCZ, and MDD). It suggested that neuroimmunity dysregulation is more severe in neurological diseases than in psychiatric disorders, led by AD. Neuroimmunity may help to redefine disease classification in the future.

Our study has several limitations. It should be noted that the disease-specific results in our study could be influenced by statistical power and sample heterogeneity. More data will be needed to develop the disease-specific signatures of neuropsychiatric disorders in the future. The difference in numbers of dIRG among disorders might be related to the sample size of each disease dataset. However, the sample size cannot explain all the differences. BD (n=188) has more samples than ASD (n=103) and PD (n=97), but has fewer dIRGs, suggesting BD involves fewer immune changes in the brain than most other disorders. In contrast, AD has possibly the most inflamed brain with more dIRGs than all other disorders, but its sample size is smaller than SCZ. We used transcriptome data from bulk tissues, which did not reflect the expression of immune genes in specific brain cell types. In our statistical procedure, we have used SVA to control hidden covariates, which minimized the cell type effects. The coexpression network analysis still suggested three cell types. Single-cell or deconvolutional data will be needed to study subtypes of cells specifically. Age and other factors may influence immune response. We have regressed out those factors when detecting dIRGs too. In order to resolve causal relationship between neuroinflammatory changes and disorders, longitudinal study will be needed.

In summary, our results provided a cross-disorder transcriptome study to explore the neuroimmune system dysfunction in six neurological and psychiatric disorders. More than 60% of the IRGs had significantly altered expression in at least one of the six disorders. The functional annotations of the dIRGs highlighted the shared dysfunction of innate immunity, and its ability to differentiate psychiatric disorders from neurological diseases. Disease-specific dIRGs and their associated pathways and coexpression modules may explain the distinct clinical features of each disorder. Stress environment may have a dominant effect on the observed changes. Our study suggests that therapeutic targeting of different components of the systems may lead to distinct effects: some are general for all brain disorders while others could help specific disorders.

## Data Availability

All data produced in the present study are available upon reasonable request to the authors

## Acknowledgments

Published microarray datasets analyzed in this study are available on Gene Expression Omnibus (accession No. GSE28521, GSE28475, GSE35978, GSE53987, GSE17612, GSE12649, GSE21138, GSE54567, GSE54568, GSE54571, GSE54572, GSE29555, GSE5281, GSE28146, GSE20168, GSE20295, GSE54282, GSE68719 and GSE11223), ArrayExpress (accession no. E-MTAB-184), or directly from the study authors.

RNA-seq data (available on Synapse with accession numbers syn4590909 and syn4587609, with access governed by NIMH Repository and Genomics Resource) were generated as part of the PsychENCODE Consortium, supported by grants U01MH103339, U01MH103365, U01MH103392, U01MH103340, U01MH103346, R01MH105472, R01MH094714, R01MH105898, R21MH102791, R21MH105881, R21MH103877, and P50MH106934 awarded to Schahram Akbarian (Icahn School of Medicine at Mount Sinai), Gregory Crawford (Duke), Stella Dracheva (Icahn School of Medicine at Mount Sinai), Peggy Farnham (USC), Mark Gerstein (Yale), Daniel Geschwind (UCLA), Thomas M. Hyde (LIBD), Andrew Jaffe (LIBD), James A. Knowles (USC), Chunyu Liu (SUNY), Dalila Pinto (Icahn School of Medicine at Mount Sinai), Nenad Sestan (Yale), Pamela Sklar (Icahn School of Medicine at Mount Sinai), Matthew State (UCSF), Patrick Sullivan (UNC), Flora Vaccarino (Yale), Sherman Weissman (Yale), Kevin White (UChicago), and Peter Zandi (JHU).

RNA-seq data from the CommonMind Consortium used in this study (Synapse accession no. syn2759792) was supported by funding from Takeda Pharmaceuticals Company, F. Hoffman-La Roche and NIH grants R01MH085542, R01MH093725, P50MH066392, P50MH080405, R01MH097276, RO1-MH-075916, P50M096891, P50MH084053S1, R37MH057881, R37MH057881S1, HHSN271201300031C, AG02219, AG05138, and MH06692.

RNA-seq data from the ROSMAP were provided by the Rush Alzheimer’s Disease Center, Rush University Medical Center, Chicago. Data collection was supported through funding by NIA grants P30AG10161 (ROS), R01AG15819 (ROSMAP; genomics and RNAseq), R01AG17917 (MAP), R01AG30146, R01AG36042 (5hC methylation, ATACseq), RC2AG036547 (H3K9Ac), R01AG36836 (RNAseq), R01AG48015 (monocyte RNAseq) RF1AG57473 (single nucleus RNAseq), U01AG32984 (genomic and whole exome sequencing), U01AG46152 (ROSMAP AMP-AD, targeted proteomics), U01AG46161(TMT proteomics), U01AG61356 (whole genome sequencing, targeted proteomics, ROSMAP AMP-AD), the Illinois Department of Public Health (ROSMAP), and the Translational Genomics Research Institute (genomic). Brain tissue for the study was obtained from the following brain bank collections: the Mount Sinai NIH Brain and Tissue Repository, the University of Pennsylvania Alzheimer’s Disease Core Center, the University of Pittsburgh NeuroBioBank and Brain and Tissue Repositories, the Harvard Brain Bank as part of the Autism Tissue Project (ATP), the Stanley Medical Research Institute, and the NIMH Human Brain Collection Core.

We thank Richard F. Kopp, Mingrui Yu, and Yan Xia for reviewing the manuscript.

## Author contributions

Conceptualization: YC, CL

Methodology: YC, JD, LT

Investigation: YC, JD, QL, QH, ML, JZ

Visualization: YC

Funding acquisition: CL, CC

Project administration: CL

Supervision: CL, CC

Writing – original draft: YC, JD, CL,

Writing – review & editing: YC, CL, JD, CC,CW

## Data and materials availability

All data are available in the main text or the supplementary materials.”

## Supplementary Materials

Materials and Methods

Supplementary Text

Figs. S1 to S8

Tables. S1 to S10

References (47-56, 95-103)

