## Supplemental Materials and Figures for "Neuroimmune transcriptome changes in brains of psychiatric and neurological disorder patients"

Data description

Twenty-three studies of multiple brain disorders were obtained from the Gene Expression Omnibus (GEO), ArrayExpress, or directly from study authors (see Supplementary Table 1). Raw microarray gene expression data was obtained from 921 postmortem cortical brain samples(microarray) and 1387 postmortem brain samples (RNA-seq). Each study was processed separately and analyzed according to the general workflow as described in Quality Control and Normalization.

Immune genes selection

Immune genes were collected from immune databases: Comparative Toxicogenomics Database(52), ImmPort(53), ImmunomeDB(54), InnateDB(55), ImmuneSigDB(56), Gene Ontology database(57), KEGG database(58), and literature reviews(59-61). The selected immune genes were included by more than two databases identified. Neuroimmunology and general immunology literature, as well as pathway annotation programs, were consulted for the selection of immune genes. Genes identified by more than two independent sources were included. We included 1,789 immune-related genes(IRGs) in our studies. 1,275 of 1,789 IRG were detected across all microarray expression data.

Supplementary Table 2 lists the immune genes categorized by their reference databases and pathway annotation programs. All IRGs were categorized into three major groups (Innate Immunity, Adaptive Immunity and Others). Because there is a high degree of overlap in a biological immune response, genes with roles in multiple immune functions were assigned to more than one gene set or group.

### Quality Control and Normalization

Known covariates included available biological (e.g., sex, age, brain region) and technical covariates (e.g., experimental batch, RIN, postmortem interval, pH) for each study from GEO. Unknown covariates were estimated by R package sva for each study(101). We used the lm function in R to adjust all the covariates (see Supplementary Figure. 1).

Outliers were defined as samples with standardized sample network connectivity Z scores < -2, as described, and were removed. Batch effects were corrected with the ComBat function of the sva package in R. Similar results were achieved using alterative batch correction methods, such as linear regression, or including batch in the final mixed effect model.

To provide a systematic nomenclature for assessment of gene expression across platforms, microarray probes were re-mapped to Ensembl gene IDs (v75; Feb 2014 data freeze) using the biomaRt package in R(102), taking the maximum mean signal across all probes available for each gene, using the collapseRows function. The collapseRows MaxMean function was explicitly developed to perform cross-platform microarray meta-analysis and has been extensively validated in its ability to increase between-study consistency and enhance reproducibility(63).

Finally, all available biological, technical, and unknown covariates except for the diagnostic group were regressed using lm function in R from each expression dataset before differential gene expression (DGE) meta-analysis.

### Differential Gene Expression Analysis

Differential gene expression analysis of microarray data was calculated using a linear mixed-effects model in the nlme package in R, with fixed effects of diagnostic group and study, and a random effect for the unique subject. This statistical framework enabled the calculation of meta-analytic beta values (FC) for each gene and disease, collapsing results from multiple studies while accounting for any subjects overlapping between studies with a random effect term. Genes were then filtered to include only those present across all studies (10,387 Ensembl gene IDs). Spearman's ρ was used to compare dIRGs meta-analysis fold change signatures across all disease pairs. Differential gene expression of RNA-seq replication data was calculated using limma with empiric Bayes moderated t-statistics, including the all the known and unknown covariates based on log2(normalized FPKM) expression values. This statistical framework enabled the calculation of logFC for each gene and disease.

Gene-Level RNAseq Replication

We evaluated the degree to which genes identified as differentially expressed in the discovery (microarray) datasets ($FDR < 0.05$) were replicated in the RNAseq data ($P < 0.05$ with concordant direction of effects), as shown in Supplementary Table 4. For comparison, we restricted our background to the 865 genes present across all microarray and RNAseq datasets (listed in Supplementary Table 1). Among these 865 genes, Supplementary Table 4 shows the number identified as differentially expressed in the discovery (microarray) dataset at $FDR < 0.05$. Among these discovery DGE genes, we identified the number that replicatedreplicate in RNAseq datasets, at $P<0.05$ and with the same direction of effect. We used Fisher's exact test to calculate odds ratios and the statistical significance of overlap between microarray DGE genes and the RNAseq replication set (all genes with $P<0.05$ and concordant direction). Regression coefficients for each gene were calculated for each group and Spearman's correlation used to assess disturbance concordance between microarray data and RNA-seq data, as above.

#### Disorder network Gene Co-Expression Network Mega-Analysis

To place results from individual genes within their systems-level network architecture, we performed Weighted Gene Co-Expression Network Analysis (WGCNA)(63). Individual (covariate-regressed) expression datasets were combined using the 10,387 genes present across all studies. ComBat was used to mitigate batch effects, as shown in sFig. 1. This normalized mega-analysis expression set was then used for all downstream network analyses.

Network analysis was performed with the WGCNA package using signed networks. A soft-threshold power of 9 was used for all studies to achieve approximate scale-free topology (R2>0.8). Networks were constructed using the blockwiseModules function. The network dendrogram was created using average linkage hierarchical clustering of the topological overlap dissimilarity matrix (1-TOM). Modules were defined as branches of the dendrogram using the hybrid dynamic tree-cutting method. Modules were summarized by their first principal component (ME, module eigengene), and modules with eigengene correlations of >0.9 were merged. Modules were color-labeled for illustration purposes. Genes that did not fall within a specific module were assigned the color grey.

Dynamic tree cut methods and signed networks were used because they are more biologically meaningful than static tree cut methods and unsigned networks. Soft threshold power was chosen to be the smallest value such that approximate scale-free topology was achieved, defined as R2 > 0.8 for the frequency.

To generate a Disease-specific network, we compared networks constructed by patients from one of seven brain disorders and networks constructed by healthy individuals separately. Then, a WGCNA integrated function (modulePreservation) was used to calculate module preservation statistics, and the Z summary score (Z score) was applied to evaluate whether a module was conserved or not. A disease-specific module is defined by z-score less than 10 in the all expression matrix of controls and other disorders patients .

Gene Set Enrichment

Gene set enrichment analysis was performed using clusterProfiler R packages(103). We used the compareCluster function to automatically calculate GO enriched functional categories of dIRGs from each disorder. GO semantic similarity can be calculated by GOSemSim. We used it to cluster genes/proteins into different clusters based on their functional similarity and also to measure the similarities among GO terms to reduce the redundancy of GO enrichment results.

Module functional enrichment of Gene Ontology pathways was assessed with GO-Elite v1.2.5(104) as well as with the gProfiler(105) R package, using GO and KEGG databases. For gProfiler, "moderate" hierarchical filtering was used. A custom background set consisted of the (10,387) genes present across all studies and microarray platforms. The top pathways reaching significance with FDR-adjusted P < 0.05 are shown in Fig. 3E and Supplementary Table 8. The two methods had highly concordant results.

Cell-type specific expression analysis of genes within each module was performed using the pSI package(106) (specificity index; http://genetics.wustl.edu/jdlab/psi_package/) in R. Cell-type specific gene expression data was obtained from an RNAseq study of purified populations of neurons, astrocytes, oligodendrocytes, microglia, and endothelial cells derived from the adult human cerebral cortex(107). Raw data (FPKM) was downloaded from GEO (GSE73721). Gene symbols were mapped to Ensembl gene identifiers using the biomaRt R package. Expression values were log2 normalized and averaged across cell-type replicates. Specificity for the five CNS cell types was calculated with the specificity.index function. Significance was assessed using Fisher's exact test with a pSI threshold set to 0.05, followed by FDR-correction of p values.

Assessment of Age Effects

To assess the influence of age on the magnitude of modules and to account for potentially non-linear effects of age, we performed a local regression analysis using the 'locfit' package in R. For each gene expression measure, a local regression function was fit to model the effect of age on expression in samples of different disorders, as follows: fit = locfit(Eigengene ~ Age, data=df[df$Group=="[Disorders group],"]). We then assessed the correlation between z-transformed expression and age within each disease group separately to identify the relationship between modules and age. The gene set enrichment analysis across brain regions and development was performed by Cell-Specific Expression Analysis (CSEA) tool[].

GWAS Enrichment

We compiled a set of GWAS summary statistics for several brain disorders, cognitive, and behavioral traits (Supplementary Table 1). Summary statistics from GWAS meta-analyses of ASD, schizophrenia, bipolar disorder, and major depression were downloaded from the PGC website (https://www.med.unc.edu/pgc/downloads). GWAS studies of educational attainment, depressive symptoms, and neuroticism were obtained from the respective studies (Table .S1).

Gene-level analysis of GWAS results was performed by MAGMA v1.04, a gene-set annotation framework that accounts for linkage disequilibrium (LD) between SNPs(108). LD was calculated using the 1000 Genomes European ancestry reference dataset. An annotation step was performed first in which SNPs were mapped to genes (hg19 genome build, depending on the study) based on the presence of a SNP in the region between a gene's start and stop sites. The gene-level analysis was then performed to create aggregate statistics for each gene. To quantify the GWAS signal enrichment within each gene coexpression module, we calculated Spearman's correlation between the module membership (kME) of each gene and the –log10 p-value for that gene for each GWAS study. kME is a measure between 0 and 1 of the centrality of a gene within a module; "hub genes" have kME values approaching 1, whereas genes that are not present in a module generally have kME less than 0.5. This process was performed for all module x GWAS combinations, and p values were FDR-corrected.

Sex comparison

To search for differences between sexes, we completed differential analysis in males and females separately. DGE was calculated using a linear mixed-effects model using the nlme package in R, with fixed effects diagnostic group and study. This statistical framework enabled the calculation of meta-analytic beta values for each gene and disease, collapsing results from multiple studies while accounting for any subjects overlapping between studies with a random effect term. Genes were then filtered to include only those present across all studies (10,387 Ensembl gene IDs). The Wilcox test was used to compare DGE meta-analysis log2FC signatures across different gender, as shown in Supplementary Table 5.

**Supplementary Figures**

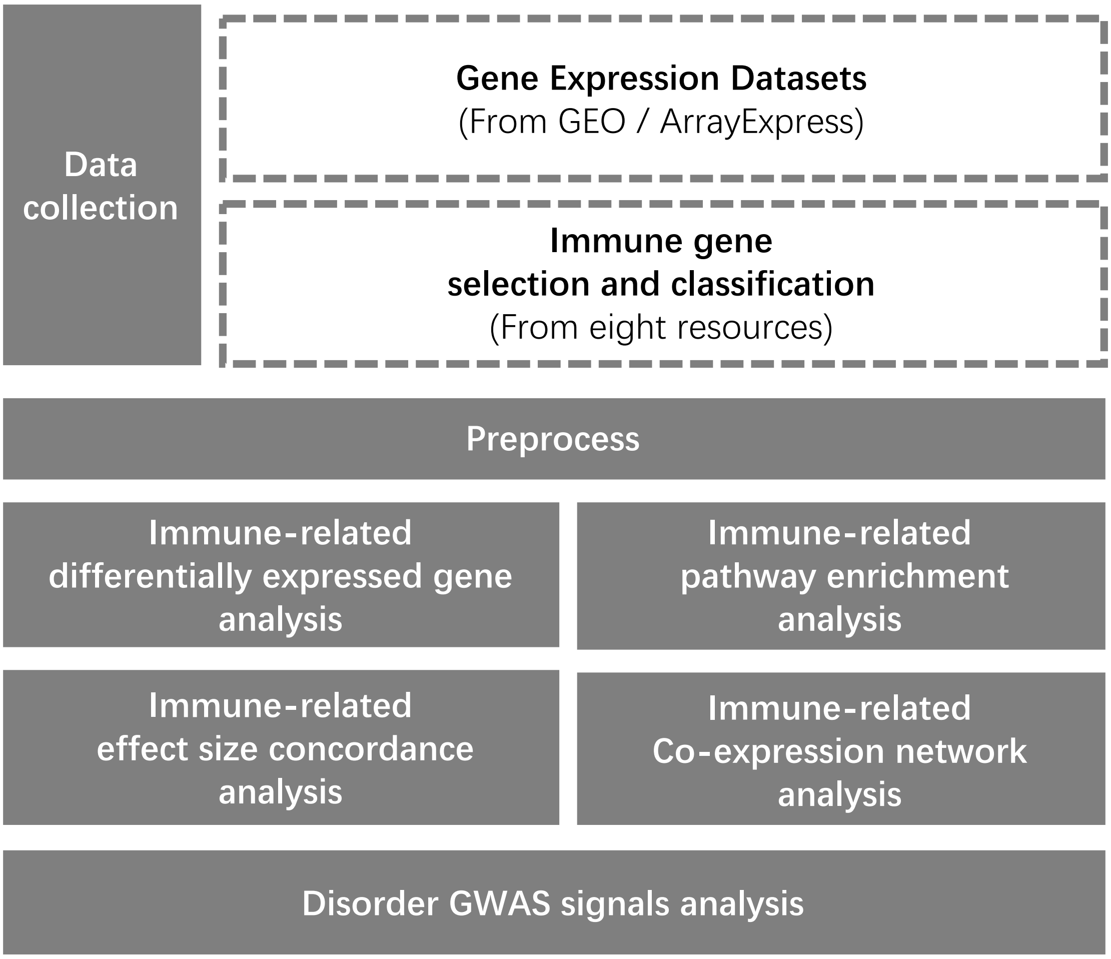


Figure S1. Study design

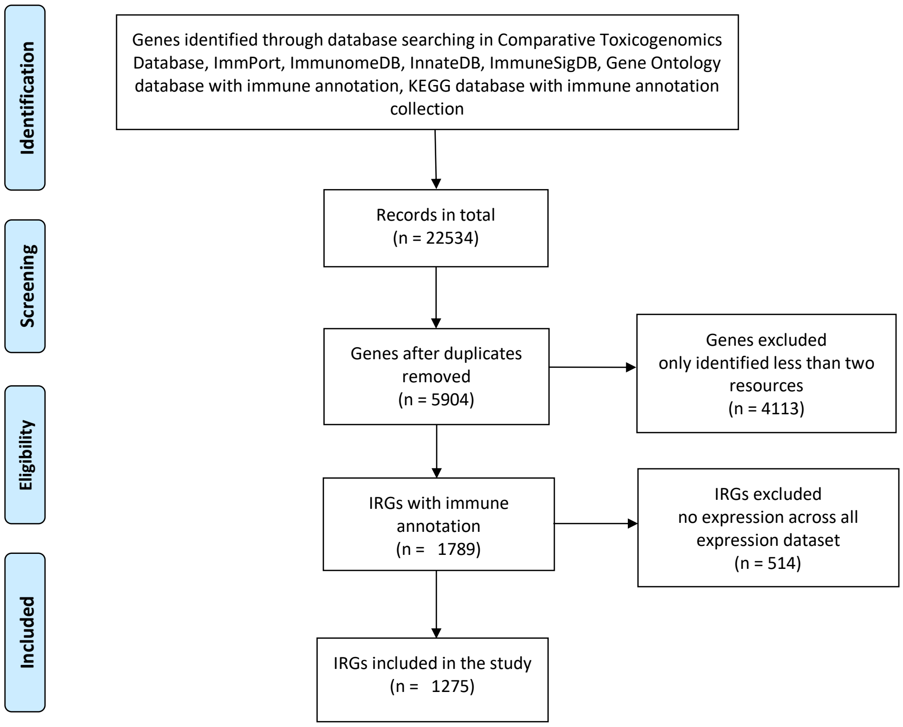


**Figure S2. PRISMA (Preferred Reporting Items for Systematic Reviews and Meta-Analyses)** It is an evidence-based minimum set of items aimed at helping authors to report a wide array of systematic reviews and meta-analyses, primarily used to assess the benefits and harms of a health care intervention. PRISMA focuses on ways in which authors can ensure a transparent and complete reporting of this type of research.

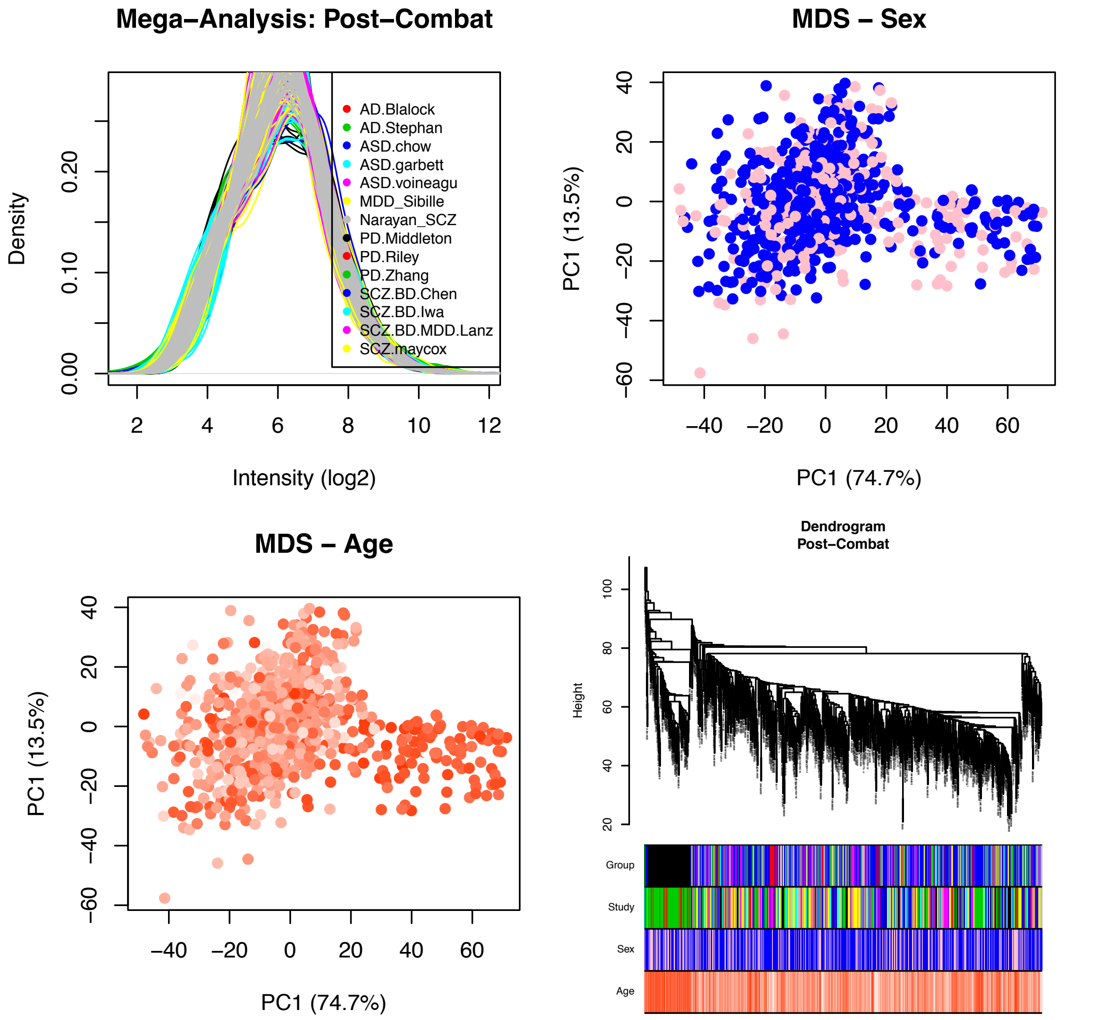


Figure S3. Here we showed post quality control plots including expression distribution and histograms. Outlier detection was determined based on standardized network connectivity z-scores. Multidimensional scaling (MDS) plots show sample clustering by the first two expression principal components.

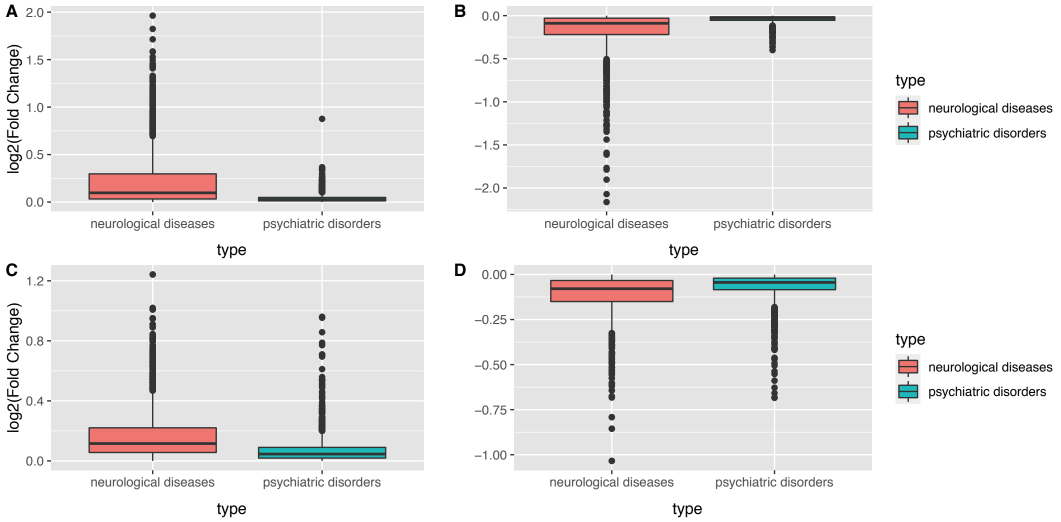


Figure S4. A, B A higher log2-FC in neurological disorders than in psychiatric disorders (t-test p.value < 2.2E-16, Fig. S4). C,D RNA-seq data replicated this observation that average log2-FC of IRGs is higher in neurological disorders than in psychiatric disorders.

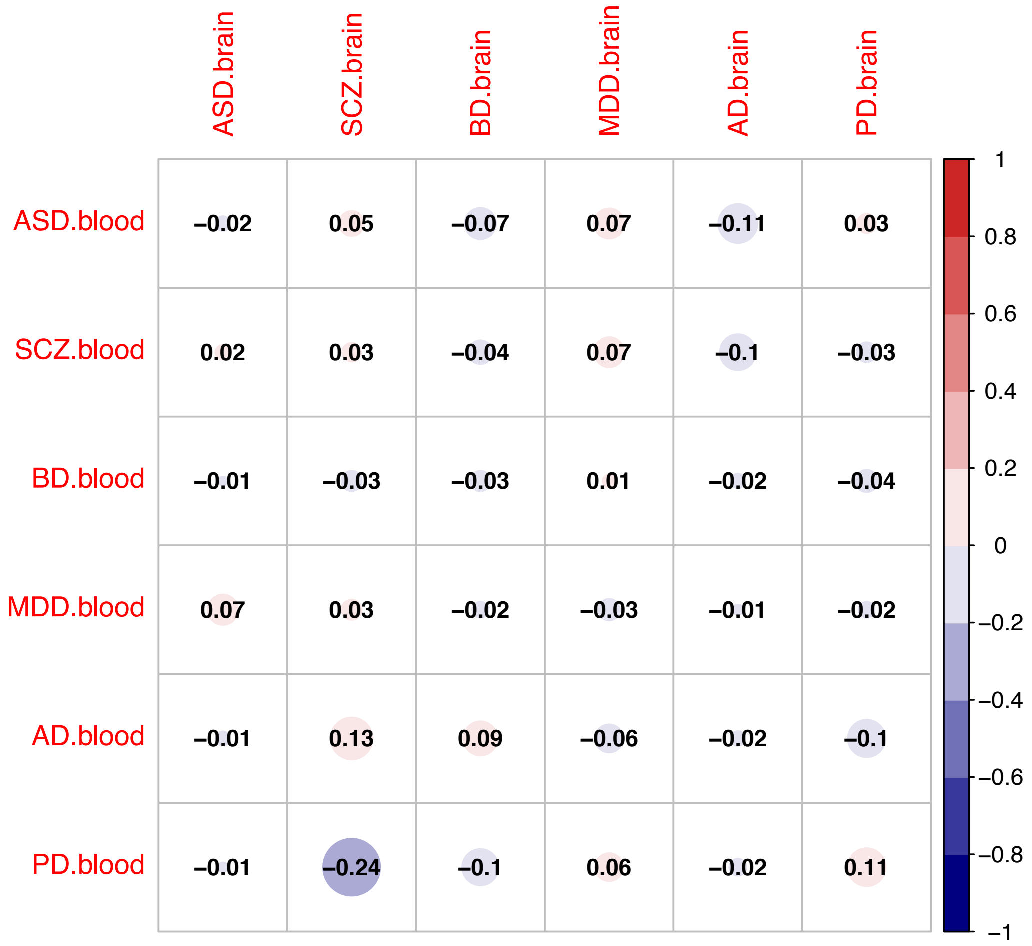


**Figure S5.** Brain blood correlations across neuropsychiatric disorders. The color of each box indicates the magnitude of the correlation. None of them were significant after Bonferroni correction.

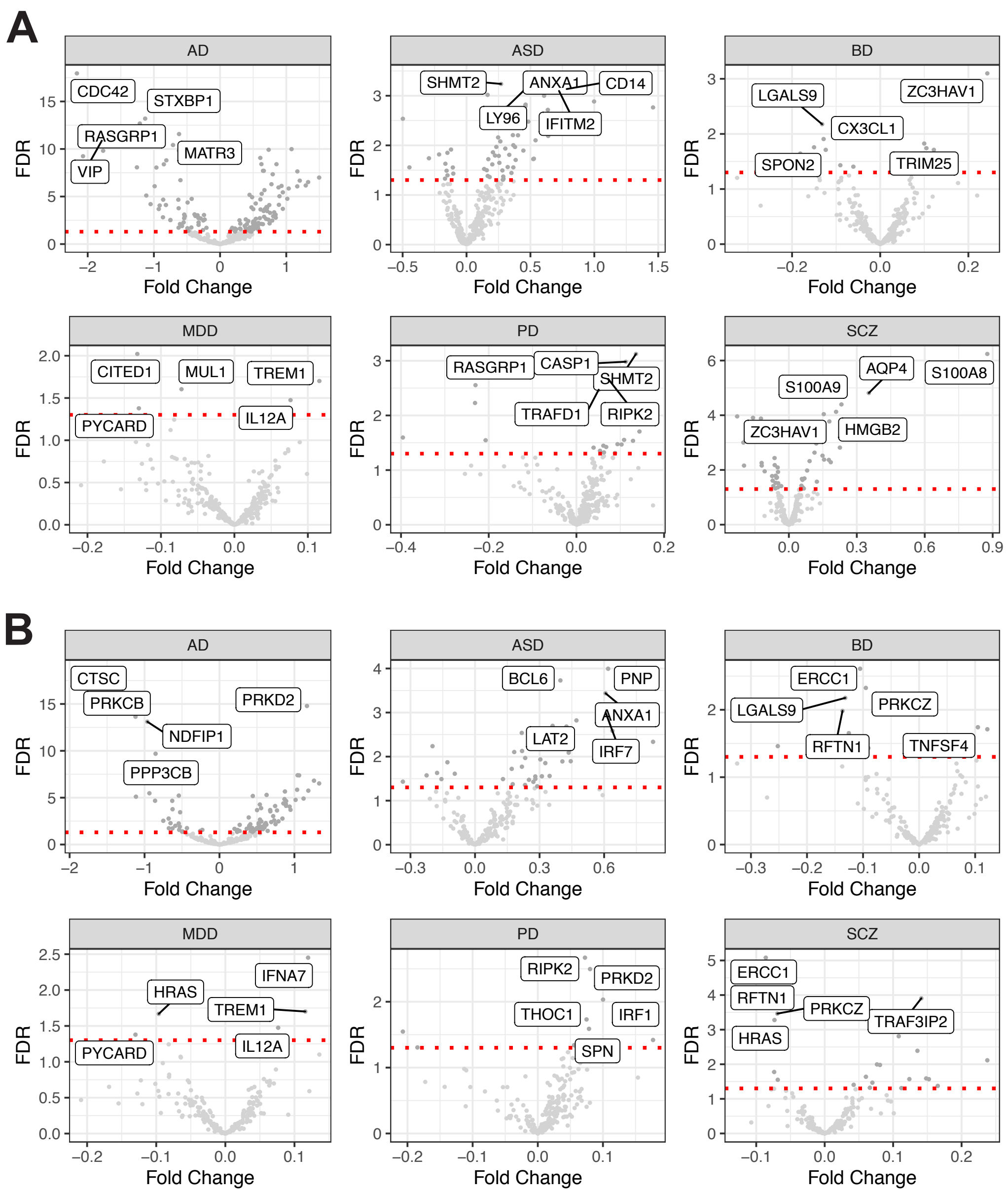


Figure S6. **Disease-specific differential expression of innate and adaptive IRGs.** A. Innate IRGs' volcano plot for each disorder. B Adaptive IRGs' volcano plots for each disorders.

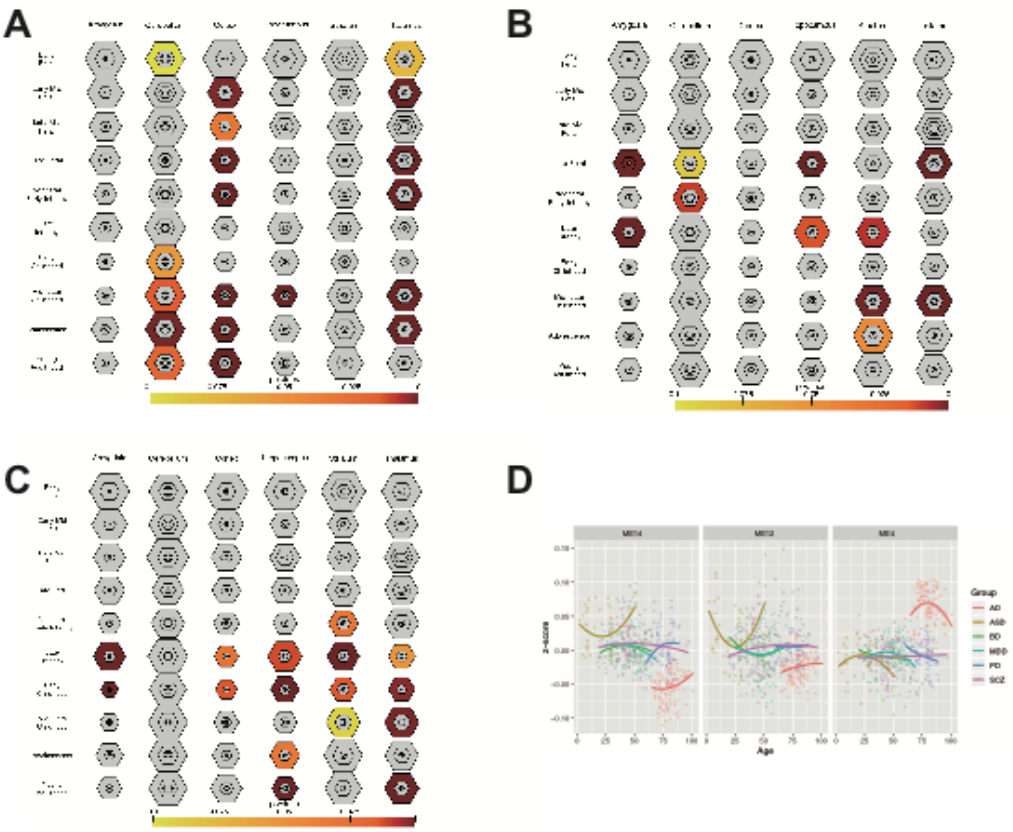


Figure S7. Analysis of enrichment in human brain regions supports the disruption of shared-IRMs. Genes of shared-IRMs are over-represented at multiple pSI levels in brain enriched transcripts calculated from Brainspan. Varying stringencies for enrichment (pSI): are represented by the size of the hexagons going from least specific lists (outer hexagons) to most specific (center). Hexagons scaled to size of gene lists.A Shared-IRM-4 has enriched expression during fetal cortical, cerebellum, and thalamus development. B. Shared-IRM-14 has enriched expression during medium-fetal brain development. C. Shared-IRM-12 has enriched expression during brain development after infancy. D. Eigengene expression of shared-IRMs across age demonstrates distinct and dynamic neural-immune trajectories.

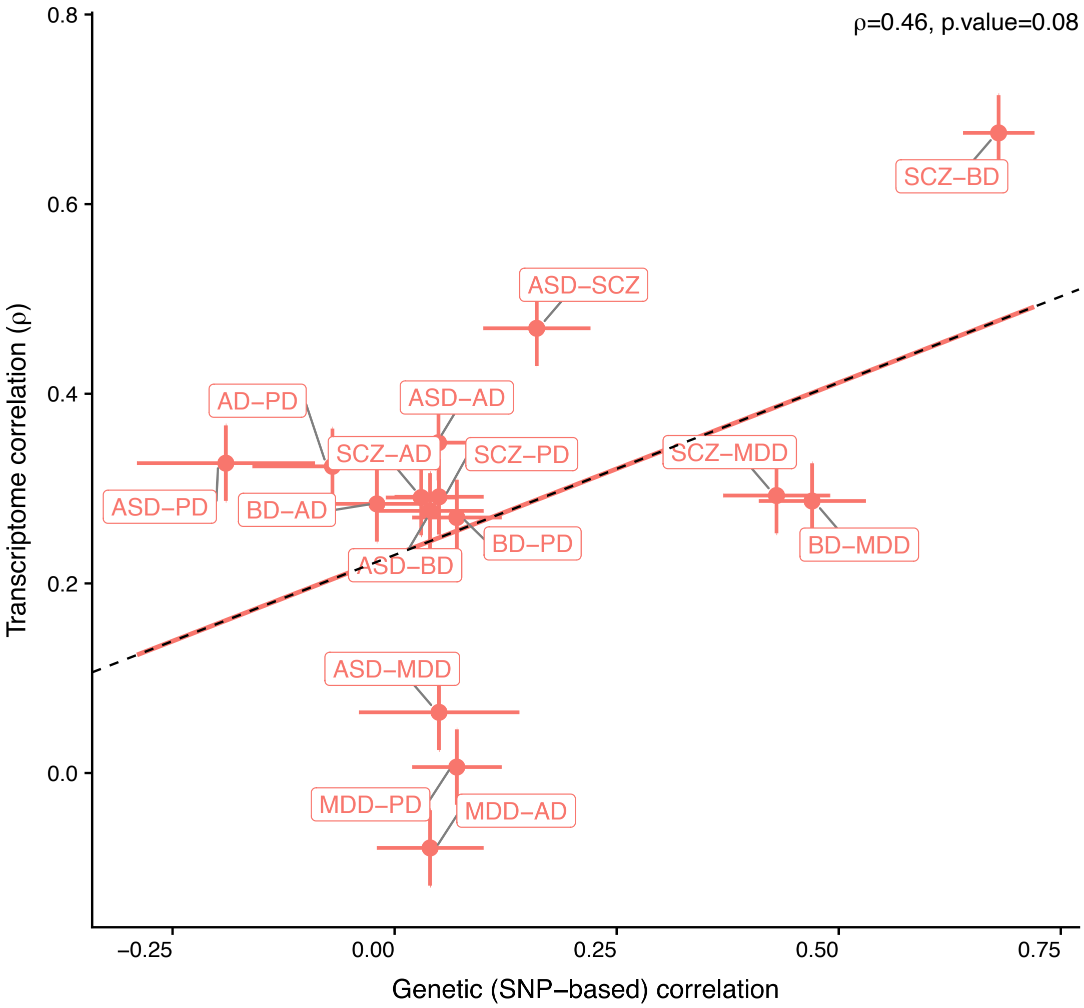


Figure S8. Correlation between transcriptome similarity and genetic overlap.
